# Identifying barriers to care and child mortality in urban and rural areas: a mixed method study in Sierra Leone

**DOI:** 10.1101/2024.10.07.24315017

**Authors:** James W.T. Elston, Kostas Danis, Nell Gray, Kim West, Kamalini Lokuge, Benjamin Black, Beverley Stringer, Augustine S. Jimmisa, Aiah Biankoe, Mohammed O. Sanko, Donald S. Kazungu, Sibylle Sang, Holly Baker, Grazia Caleo

**Author notes:** **Corresponding author:** Dr Grazia Caleo. Author qualifications (selected): Dr James Elston MBBS MRCP MFPH CertMedEd DTM&H DLSTM MSc Kostas Danis MD MSc PhD Nell Gray MA MA Kim West BSc MPH Grazia Caleo MD MSc DTM&H.

## Abstract

**Background:** Reducing mortality of children <5 years in Sierra Leone is a priority. Despite an enabling policy environment, health indicators have remained poor. Evidence on barriers to care is limited.

**Objectives:** This study describes barriers to care, health-seeking behaviour, and health outcomes of children <5 years.

**Methods:** From October 2016 to January 2017, we conducted a sequential mixed-methods study in urban and rural areas of Tonkolili District comprising: household survey targeting carers of children <5 years; and in-depth interviews (IDIs) targeting community leaders and healthcare workers (HCWs). We chose 30 clusters in urban and 30 clusters in rural areas. Topics that were identified during the survey were examined further through IDIs.

**Results:** We surveyed 643 carers of 1092 children <5 years and conducted 72 IDIs. Of children <5 years, 62% had experienced febrile illness in the 2 weeks prior, and mortality was higher rurally (1.55/10,000/day vs urban 0.26/10,000/day). Barriers, including costs and physical inaccessibility of healthcare facilities, delayed or prevented 90% (287/318; 95%CI: 80-96) rural and 48% (155/325; 95%CI: 37-58) urban carers from accessing care for a febrile child. Mistrust of HCWs was frequent, primarily due to their requests for payment. HCWs described lack of pay and holistic support precluding provision of quality care.

**Conclusions:** Children <5 faced important barriers to healthcare, particularly in rural areas, contributing to high preventable mortality near to the emergency threshold. Access to healthcare was important to carers, however available services were costly and unreachable. Equally, HCWs experienced structural barriers to provide quality care.

**Key messages:**

- Child <5years mortality near to the humanitarian emergency threshold and substantially high among rural children.
- Inequity in healthcare access and inequality in health between urban and rural areas.
- Barriers, including costs of healthcare and physical inaccessibility of healthcare facilities, delayed or prevented 90% of rural and 48% of urban carers from accessing care for a febrile child.
- Just 8% of rural children <5 years used Long-Lasting Insecticide-Treated bednets.
- Mistrust of healthcare workers was widely expressed primarily due to payment demanded for “free” healthcare. Healthcare workers described lack of pay and poor conditions precluding provision of quality care.

## Background

Sierra Leone is one of the poorest countries in the world, ranking 182nd out of 189 countries in the Human Development Index in 2020.[1] The population has amongst the highest reported mortality rates for children under 5 years of age worldwide.[2] The vast majority of these deaths are preventable, with infections, in particular malaria, the leading cause of death.[3] To improve access to care, the Sierra Leonean government introduced the Free Health Care Initiative (FHCI) in 2010 mandating free provision of healthcare for children under 5 years.[4] Reducing child mortality has been a long-term priority for health strategy.[5,6,7,8] However, despite an enabling policy environment, health indicators have remained poor. Furthermore, the 2014-2016 Ebola outbreak resulted in: a substantial number of healthcare worker deaths; reduced services including vaccination coverage; a change in health seeking behaviour at population level with reduced attendances to health facilities among febrile children under 5 years; all of this leading to health consequences probably worse than those attributed to Ebola.[9,10,11,12,13,14,15] Post-Ebola, evidence for health status and health seeking behaviour for children remain lacking.

Delay in receiving adequate care during febrile illness is perceived to be a major contributor to morbidity and mortality in children.[16] The “three delays” model (delay in: i) deciding to seek care, ii) in reaching care, and iii) in receiving adequate healthcare), was developed to better understand contributors to high maternal mortality in resource poor settings, though can be similarly applied to child mortality.[16]

Médecins Sans Frontières (MSF) has been providing maternal and child healthcare in Tonkolili District since January 2016, in collaboration with the MoHS. The aim of this study was to describe health of children under 5 years, exploring health-seeking behaviours and barriers to healthcare, using the “three delays” model in a rural and urban area of Sierra Leone since the Ebola outbreak. The findings will inform health policy and service planning.

## Methods

### Setting

The study was conducted in Tonkolili District. Tonkolili (area∼7003 km^2^) is located in the centre of the country and has a mostly rural population of ∼435,000 (2004 census projections). Most of the population is of Temne ethnicity, however there are also minority groups of Koranko, Kono and Limba ethnicity. The capital and largest city is Magburaka. Tonkolili is an under-resourced district with a poor road network that floods during rainy season, causing many areas to become inaccessible. During the Ebola outbreak, the district reported 406 EVD cases that included 162 deaths.[17,18] This study was implemented separately in Magburaka town, hereby referred to as ‘urban area(s)’; and Yoni chiefdom, hereby referred to as ‘rural area(s)’.

### Study design and data collection

A mixed methods sequential explanatory design was employed, consisting of two distinct phases conducted in urban and rural areas. Phase 1 was quantitative, comprising a household survey.

Survey questionnaires were carried out with caregivers of children under 5 years. In the household survey, information on health seeking behaviour, barriers to healthcare, and child mortality was collected, although information regarding cause of death was not collected. Mortality estimation included children who were live born but died from any cause during the two years preceding the day of the survey and were aged under 5 years at the time of death. In Phase 2, qualitative in-depth interviews (IDIs) explored barriers and enablers to access care. Implementation of Phase 2 was informed by the findings of Phase 1. This study was performed concurrently with a maternal health survey, the result of which have been published.[19]

#### Household survey (Phase 1)

From October to November 2016, we conducted the household survey using a two-stage cluster sampling methodology, with minimum target sample size of 450 children under 5 years in each of two areas (rural and urban), assuming a design effect of 4 and that almost all children under 5 years in this setting would have experienced fever in a three-month period.[7] Thirty clusters were selected from each area, so we aimed to get information on 15 children of under 5 years per cluster. We estimated that ten caregivers per cluster were sufficient to provide this information. This was calculated using population estimates derived from national census projections and household composition from the Demographic and Health Survey (DHS) 2013.[7] The main outcome was utilisation of health facilities by children aged under 5 years during their most recent febrile illness in the preceding three months (assumption, from extrapolation of previous survey results, being that almost all children under 5 years in this setting would have experienced fever in a three-month period.[7] The 190 women in each of the two areas that were included in the concurrent maternal survey were additionally asked their birth history to estimate infant mortality for the purposes of this study.[19]

Cluster, household and individual sampling methodology was as per the concurrent maternal health survey.[19] Clusters were sampled by selection of random GPS points in the urban area, and by random village selection, stratified by population size, (either ≥500 or <500), in the rural area. Survey participants were i) caregivers of children under 5 years within the selected household and ii) women who had given birth in Tonkolili District since the Ebola outbreak was declared in Sierra Leone in mid-May 2014 (these women were participants of the concurrent maternal health survey).[19]

A maximum of one eligible caregiver and one eligible woman per household was included; in households with more than one eligible carer or woman, lots were drawn to determine who would participate. A number of carers sufficient to provide information relating to 15 children under 5 years and ten women were recruited for each cluster. Bespoke questionnaires were used to collect data on household demographics, use of malaria bed nets, health behaviour during most recent febrile illness, barriers to healthcare and whether these barriers impacted access to quality health care; child health outcome, and vaccination status (carers were asked to provide vaccination cards if available). Most recent febrile illness was defined as ‘illness with fever (an abnormally high body temperature usually accompanied by shivering)’ that had affected a child under 5 years within the preceding 3 months (from the day of the survey). Health behaviours were collected on children who had experienced febrile illness, however barriers to healthcare were asked only once of the carer and was concerning the most recent episode of febrile illness of the children under their care. Women were asked about the status of all children born within the past eight years, to identify all children who had been under 5 years of age at any point during the study period. Questionnaires were uploaded to Sony Experia tablets in English and administered verbally in the local language (Temne, Mende or Krio) by trained data collectors. Responses were entered into Dharma platform software by the survey team, that was of comprised eight data collection teams (two people in each team) and two to four supervisors per day. Piloting and training of the survey team was conducted.[19]

#### In-depth interviews (Phase 2)

From December 2016 to January 2017, IDIs were conducted with caregivers for a child under 5 years; women who had a live birth or stillbirth since the start of the Ebola outbreak in Sierra Leone; community leaders; and skilled or unskilled healthcare workers (HCW).

Selection of locations for IDIs was purposive, informed by preliminary analysis of the survey data to target those with the highest frequency and severity of barriers, and to include a broad range of characteristics (i.e. good/poor access to a health facility; on a main road/remote).[19]

The required sample size was established as the study progressed;[20] interviews were conducted until theoretical saturation.[21,22]

Participants were selected purposively. Maximum variation sampling was used. Topic guides were piloted and IDIs conducted, translated, transcribed and quality controlled.[19]

Interviews were conducted by one researcher and one research assistant. The IDI teams also included transcribers.

### Data analysis

Means or medians (range) were calculated for numerical variable and proportions were calculated for categorical variables, using the non-missing values as denominators and 95% confidence intervals (95%CI) allowing for clustering. Adjusted prevalence ratios (PR) were calculated using Poisson regression to assess potential associations. STATA v14 (Stata Corporation, Texas, USA) was used for data cleaning and data analysis.

Open coding was conducted to identify any emerging phenomena and patterns emerging from IDIs. Codes were grouped as per the three delays model, whilst also allowing for the identification of categories/themes. Variations across sub-groups were assessed. All irregular cases were analysed to confirm the emerging patterns. Qualitative data was analysed using NVivo ©11 software.

## Results

### Overview

In urban areas, information on 529 children under 5 years was provided by 325 carers; 301 women who had given birth since Ebola were also included. In rural areas, 318 carers provided information on 563 children under 5 years; 307 women who had given birth since Ebola were included (Table 1).

**Table 1:** Overview of household and study participants by place.

| Study component | Participant group | Number included |  |  |
| --- | --- | --- | --- | --- |
|  |  | Urban | Rural | Total |
| Survey | Households | 386 | 356 | 742 |
|  | Carers of children <5 years | 325 | 318 | 643 |
|  | Children <5 years* | 529 | 563 | 1092 |
|  | Women (given birth since Ebola) | 301 | 307 | 608 |
| In-depth interviews | Mothers | 9 | 10 | 19 |
|  | Caregivers | 6 | 6 | 12 |
|  | Community leaders | 8 | 9 | 17 |
|  | Health workers | 12 | 12 | 24 |

\*Information provided by carer

72 IDIs were conducted in survey cluster sites: 5 urban locations and 6 rural villages (Table 1). More interviews were conducted with HCWs compared to other groups as they represented the most heterogeneous group (Table 1). No refusals to participate were reported.

Household demographics in terms of household size and age were similar in the two areas (table 1 – report difference only): Rural carers overall had lower levels of literacy and lower educational attainment and were more likely to be married (Table 2). The age of women (providing information for child mortality) was also similar: median 25 years urban vs 26 years rurally; their demographics are described in detail elsewhere.[19] IDI participants were aged between 18 and 40 years; of these, 15 (79%) were aged between 18 and 30.

**Table 2:** Demographics of carers and children <5 years included in survey.

| Participant group | Characteristic | Variable | Urban |  |  |  | Rural |  |  |  |
| --- | --- | --- | --- | --- | --- | --- | --- | --- | --- | --- |
|  |  |  | # | Total | % | 95%CI | # | Total | % | 95%CI |
| Carers of children<br><5years | Sex | Female | 315 | 325 | 97% | 94-98 | 307 | 318 | 97% | 94-98 |
|  | Literacy and<br>education | Literate (able to read and<br>write) | 164 | 325 | 50% | 43-58 | 49 | 318 | 15% | 9.7-24 |
|  |  | Educated to primary<br>level | 160 | 325 | 49% | 42-57 | 251 | 318 | 79% | 70-86 |
|  |  | Educated to secondary<br>level | 21 | 325 | 6.5% | 3.8-11 | 28 | 318 | 8.8% | 5.4-14 |
|  |  | Educated to higher level | 126 | 325 | 39% | 33-45 | 39 | 318 | 12% | 7.8-19 |
|  | Marital status | Never married | 65 | 325 | 20% | 16-25 | 19 | 318 | 6.0% | 3.1-11 |
|  |  | Married | 245 | 325 | 75% | 69-81 | 285 | 318 | 90% | 85-93 |
|  |  | Divorced | 1 | 325 | 0.3% | 0.0-2.3 | 2 | 318 | 0.6% | 0.2-2.6 |
|  |  | Widowed | 13 | 325 | 4.0% | 2.2-7.1 | 12 | 318 | 3.8% | 2.1-6.6 |
|  | Relationship to child | Parent | 435 | 529 | 82% | 75-88 | 469 | 563 | 83% | 71-91 |
|  |  | Grandparent | 44 | 529 | 8.3% | 5.5-13 | 29 | 563 | 5.2% | 3.0-8.7 |
|  |  | Sibling | 4 | 529 | 0.8% | 0.3-2.0 | 12 | 563 | 2.1% | 0.7-6.5 |
|  |  | Aunt/uncle/cousin | 39 | 529 | 7.4% | 3.6-15 | 38 | 563 | 6.7% | 3.7-12 |
|  |  | non-related | 7 | 529 | 1.3% | 0.5-3.5 | 15 | 563 | 2.7% | 0.8-8.2 |
| Child <5years | Sex | Female | 245 | 529 | 46% | 42-51 | 306 | 563 | 54% | 51-58 |

### Health outcomes

Overall, 62% of children under 5 years had experienced febrile illness in the 2 weeks prior to the day of the survey, and 90% within the preceding 3 months (Table 3). Whilst the frequency of illness was similar between areas, almost all urban children had recovered from their most recent febrile illness on the day of the survey, whilst 14% of rural children had shown no signs of improvement or had got worse (Table 3). 17 (3%) of live born urban children had died in the preceding 8-year period compared with 122 (16%) of rural children (Table 3). The under 5 years mortality rate in the urban area was 0.26/10,000/day (95%CI: 0.16-0.43) vs 1.55/10,000/day (95%CI: 1.30-1.86) rurally.

**Table 3:**
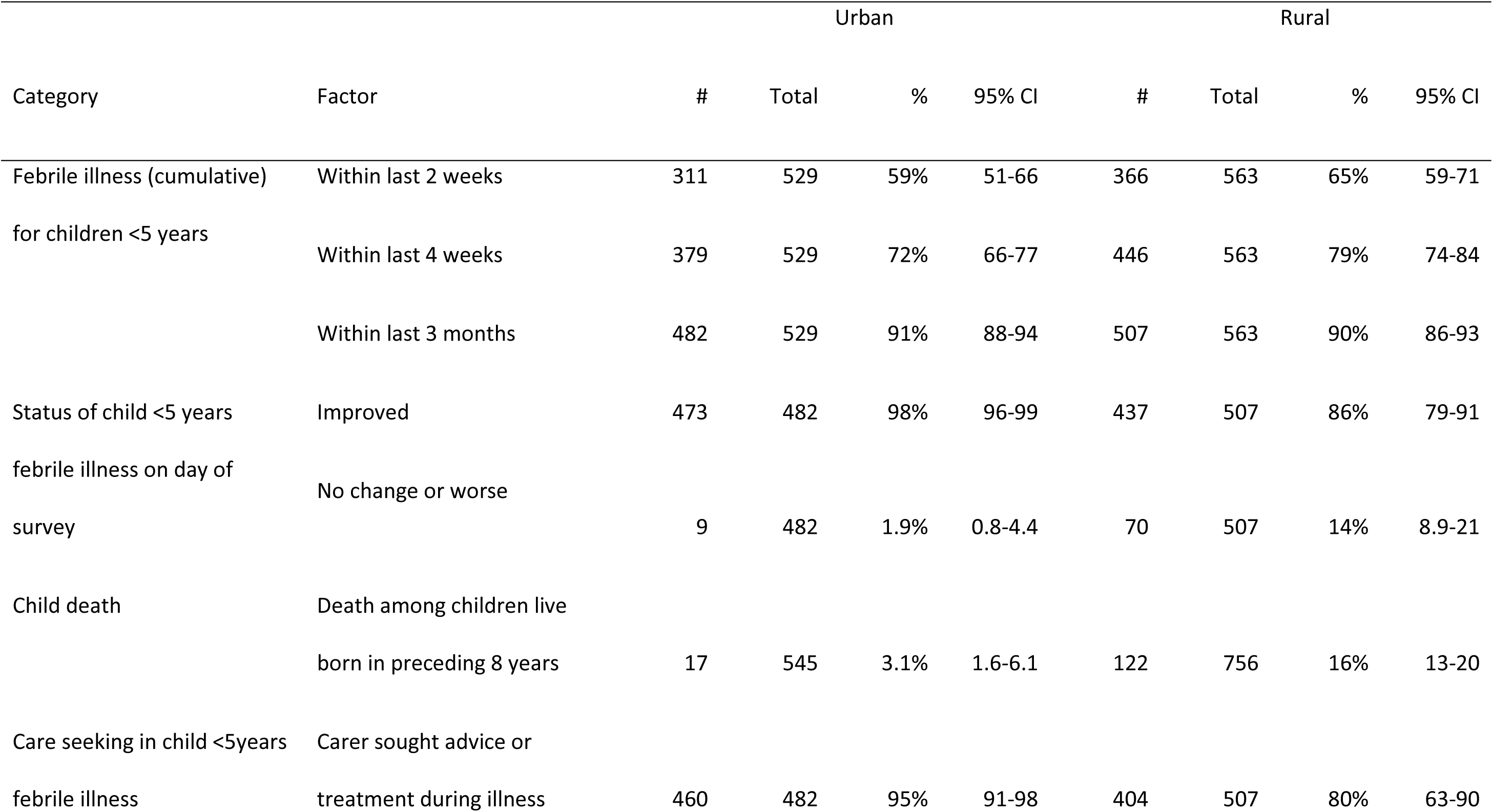
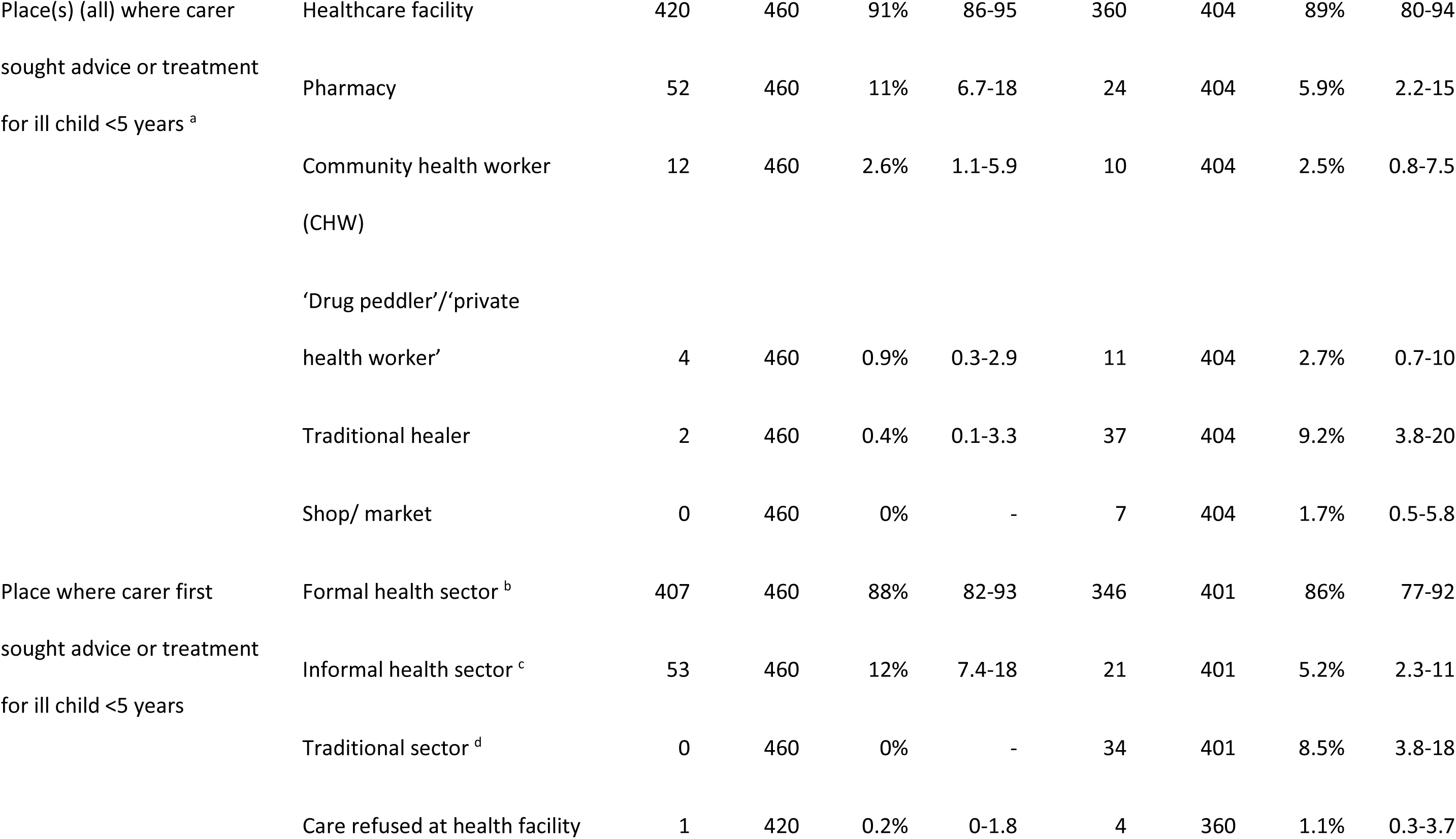
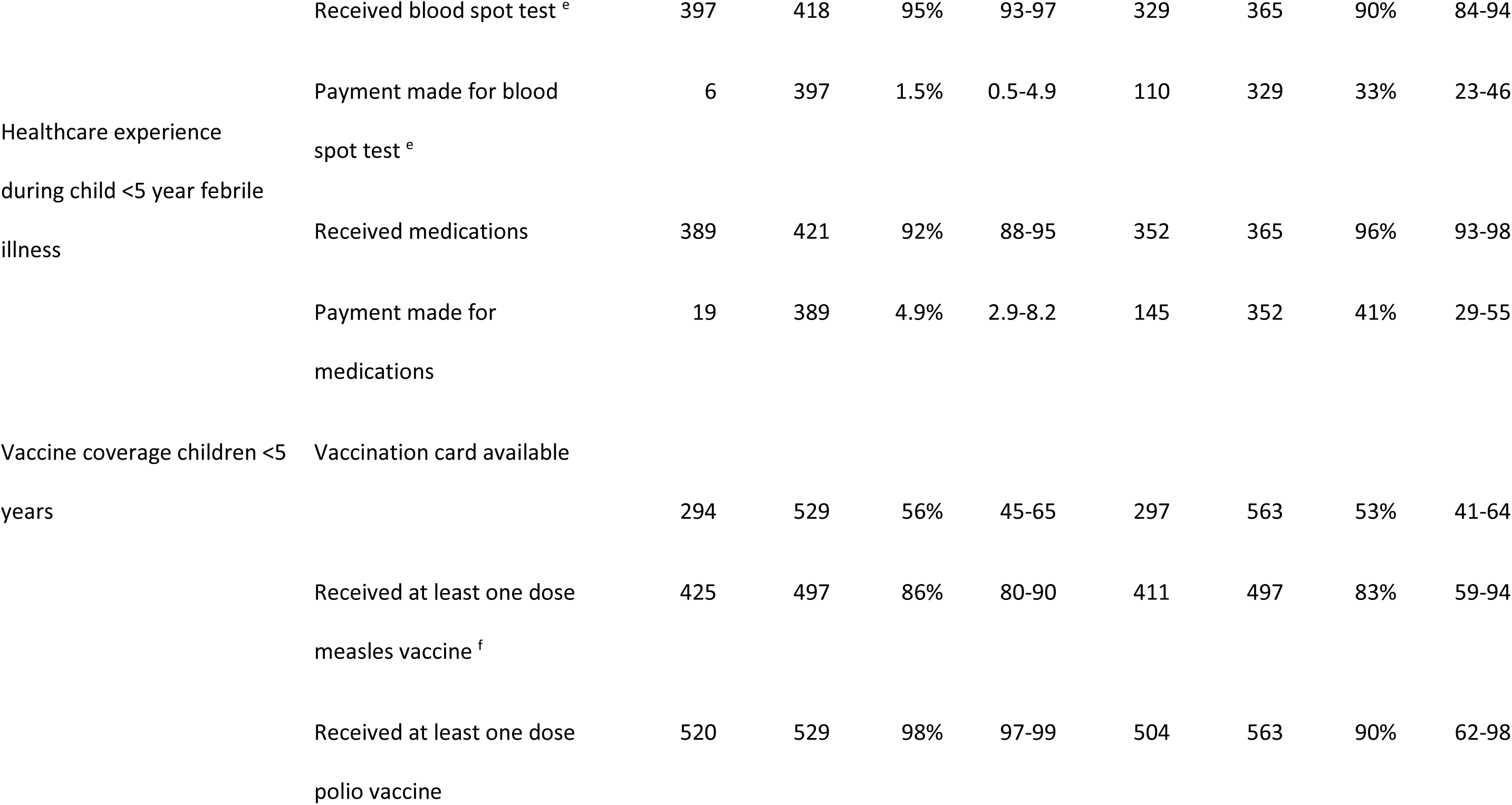
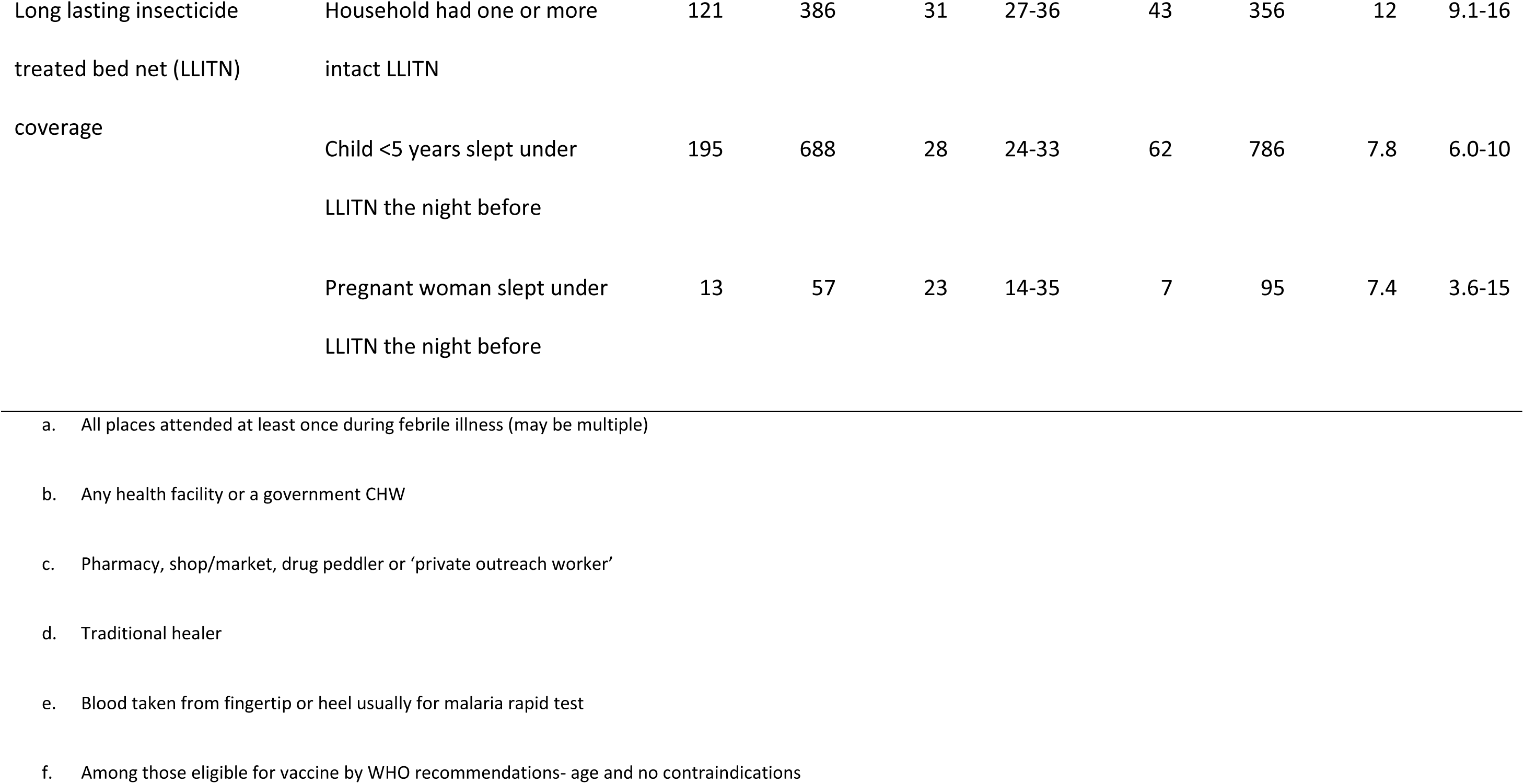
Health outcomes, health behaviours, healthcare experiences and vaccine coverage for children <5years, and household LLITN coverage.

### Barriers to healthcare

48% (155/325; 95%CI: 37-58) of urban and 90% (287/318; 95%CI: 80-96) of rural carers experienced at least one problem which delayed or prevented them from being able to receive healthcare for a febrile child under 5 years under their care, at any stage during the most recent illness. Rural carers generally reported a greater number of barriers, and attendances for children under their care were delayed and prevented to a more substantial degree compared with urban carers (Figure 1). Lack of money to pay for a consultation and/or transport was reported as a barrier for 32% (95%CI: 24-41) of urban carers and 86% (95%CI: 76-93) of rural carers. A substantial proportion of carers, especially in the rural areas, cited distance to a health facility, fear of contracting Ebola, not wanting to travel alone and concerns about being treated disrespectfully by HCWs as reasons for delaying or abandoning their attempt to access healthcare (Figure 1).

**Figure 1:**
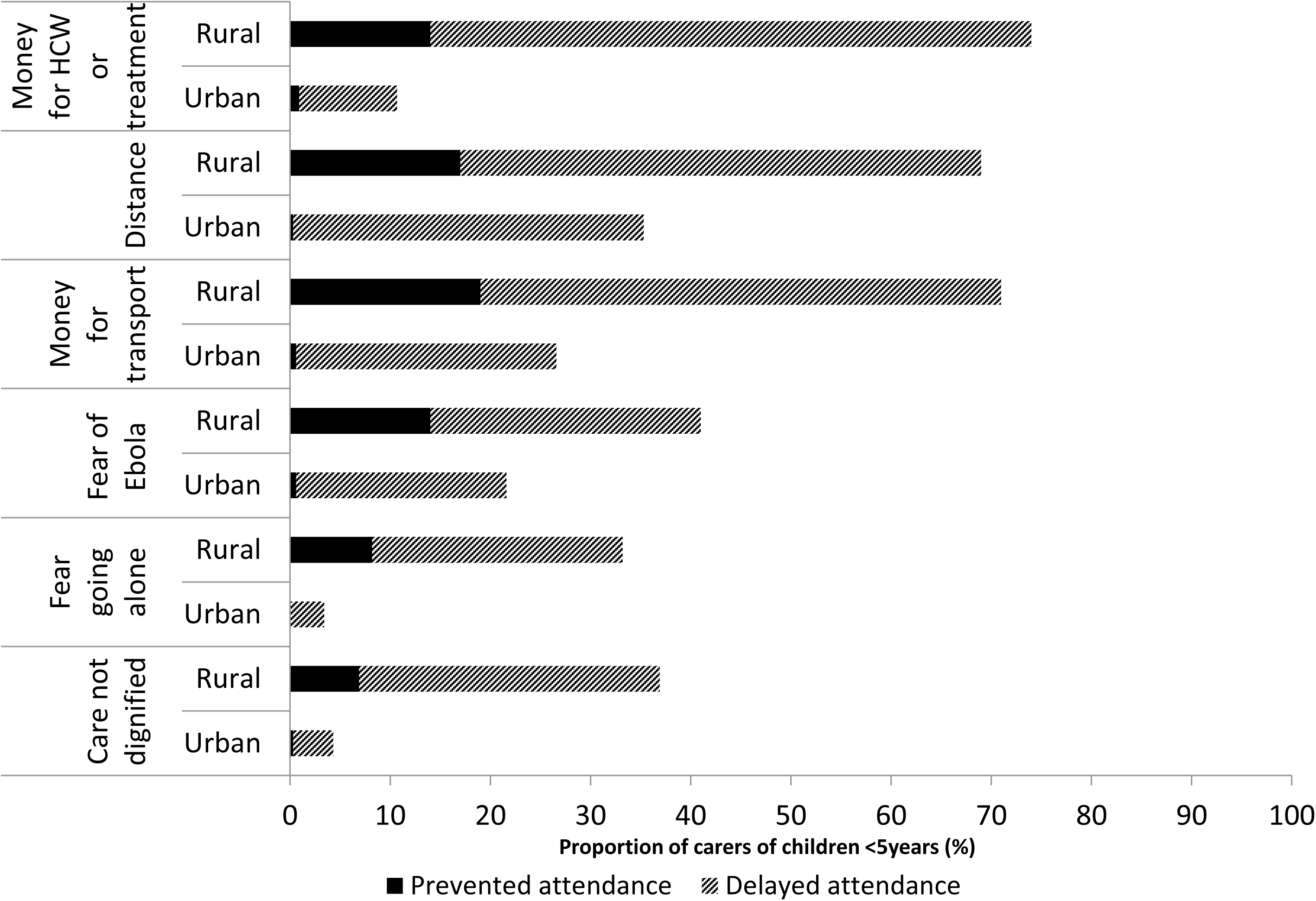
Factors delaying or preventing carers from seeking/ reaching healthcare for febrile children <5 years by place

Lack of medications and HCW absences were also reported as barriers.

### Care seeking and delays in deciding to seek care

95% of urban vs 80% of rural carers sought advice or treatment (from any source) when the child was unwell with their (most recent) febrile illness (Table 3).

In the rural area, more educated carers were more likely to have sought advice/ treatment for their febrile child but those who cared for more children were less likely to have done so: the proportion increased by 10% for each increasing level of the carers educational attainment but decreased by 14% for each additional child they cared for (Table 4). No such associations were identified for urban carers (Table 4).

**Table 4:**
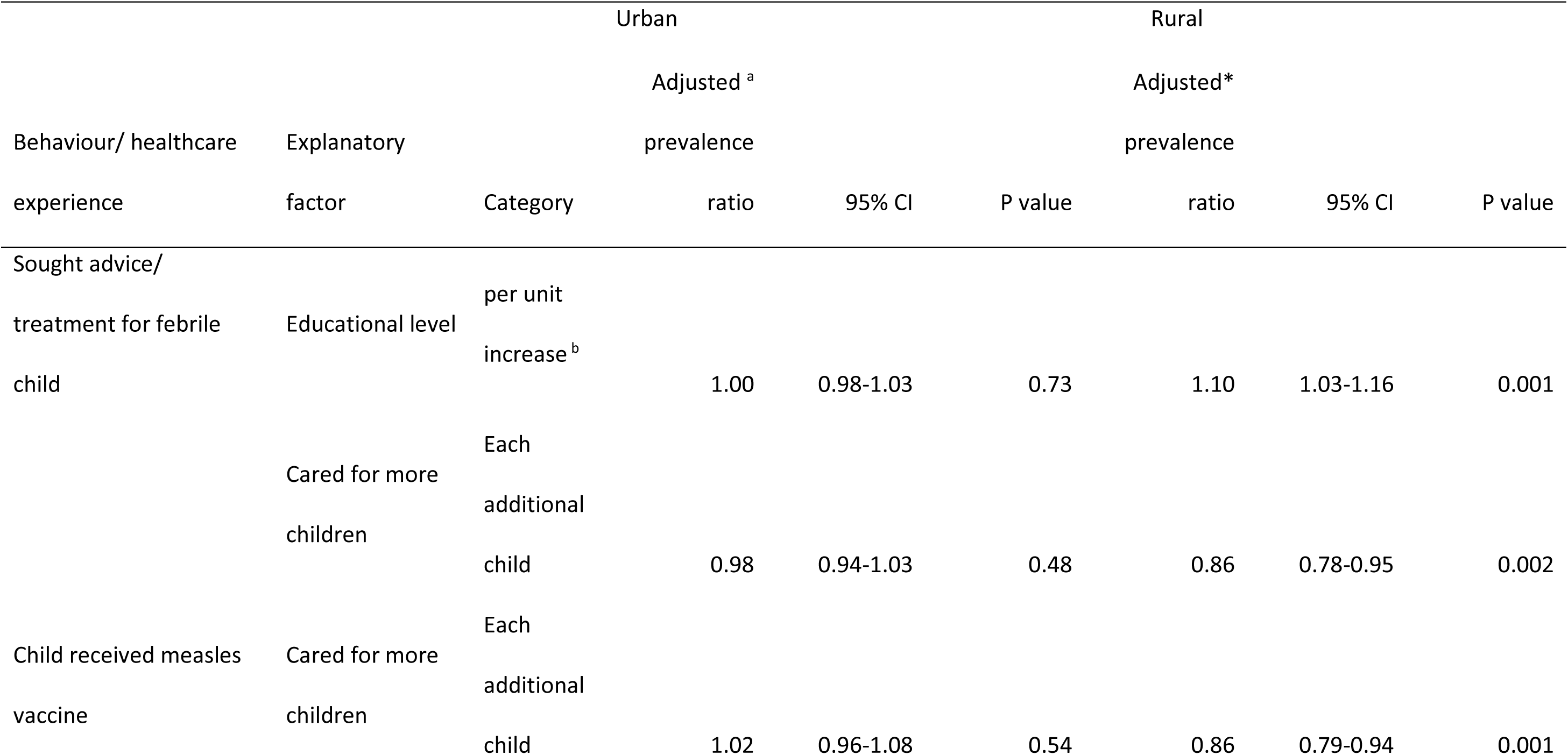
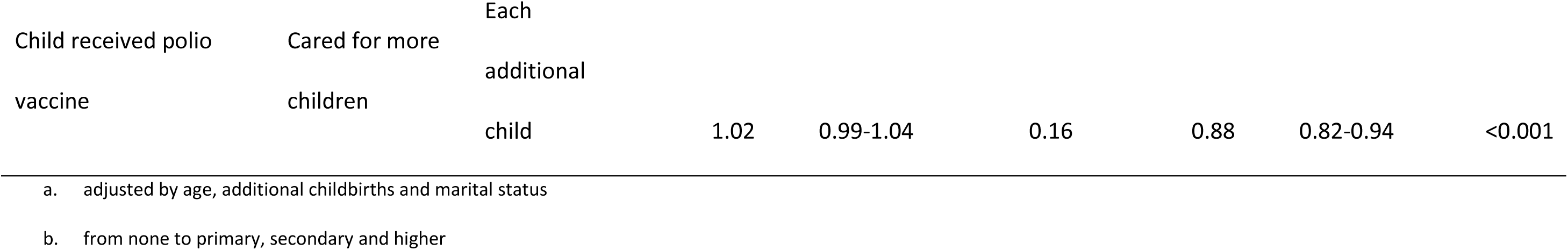
Demographic associations with health seeking behaviour.

In both urban and rural areas, IDI participants reported the main factors for delaying care-seeking were concerns about prohibitive costs associated with seeking care and previous experiences of poor care (e.g. described as being treated disrespectfully by HCWs and not being given perceived necessary medication) (Box 1). Additionally, care seeking was delayed by concerns about long waiting times in urban areas, and distance to health facilities, lack of transport options, poor roads, and absent staff in rural areas. Mothers also described the difficult decisions they faced in allocating scarce resources (money and time) to attend a health facility for a sick child in the face of competing responsibilities to care for large families and to generate income.

**Box 1:**
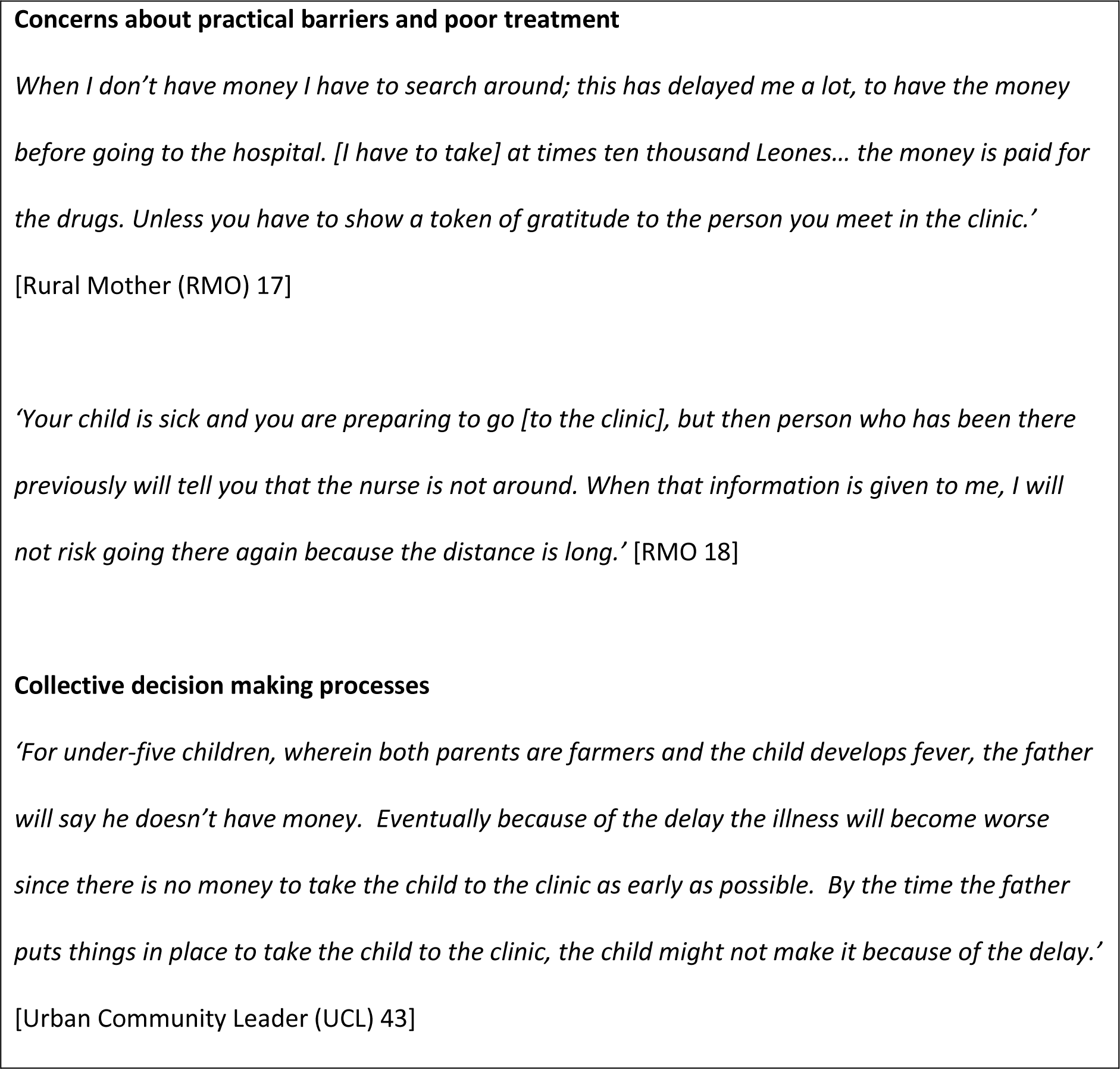
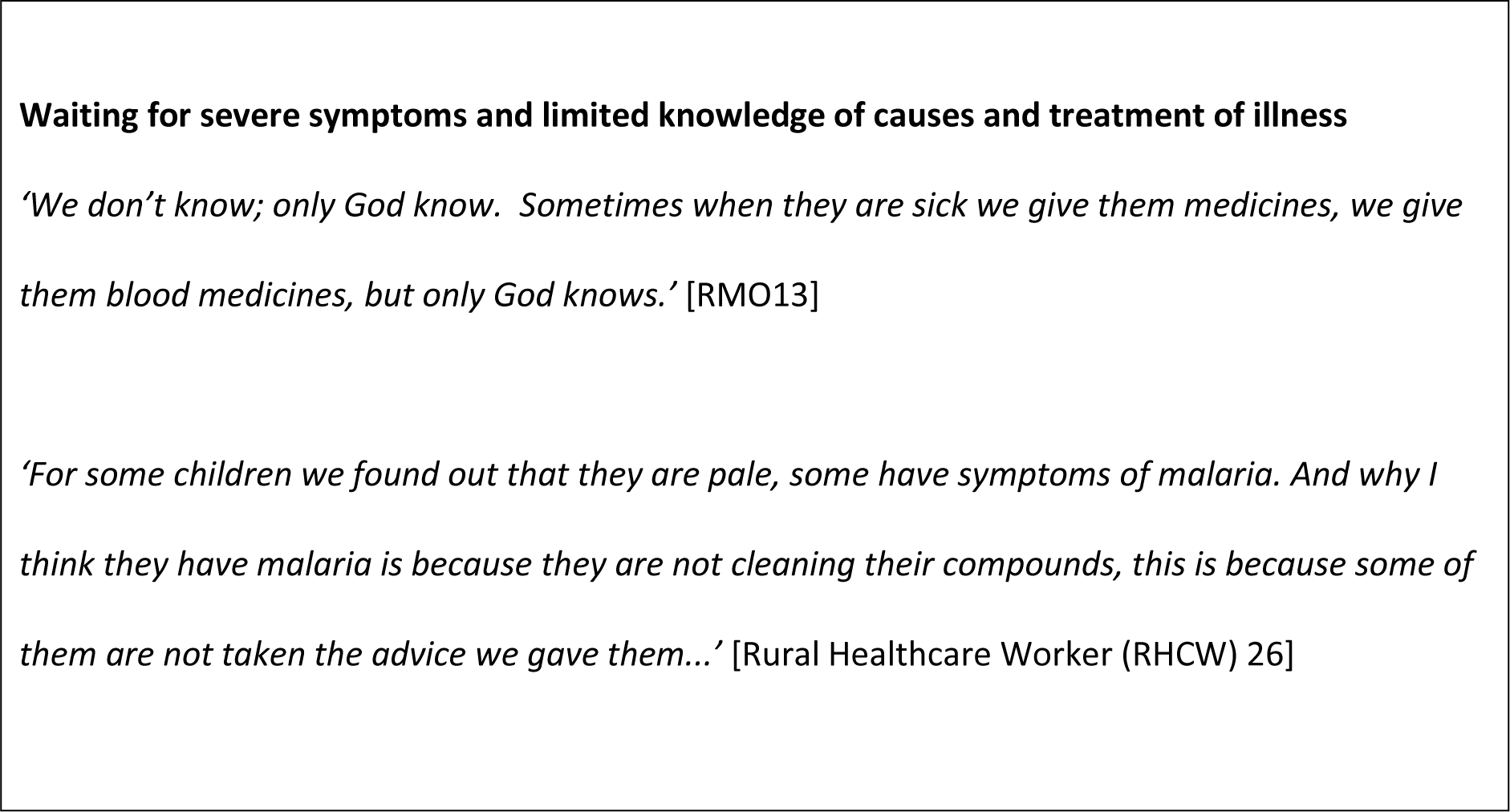
Themes and quotes relating to delayed care seeking.

Both rural and urban IDI participants reported time to reach a consensus within the family on choice of care provider as a delaying factor (Box 1). Husbands played a critical role in decision making as they generally controlled family finances. Additionally, participants reported that extended family members (notably mothers and mothers-in-law) as well as influential community members frequently advised on appropriate care provision. Notably, several participants indicated that the ‘older generation’ often advocated for ‘traditional’ care whereas peers and health messaging promoted attendance of formal services, and that managing this varied and often contradictory advice was challenging. Faith also played a role in care seeking behaviour, with several participants explaining that the life or death of their child was in the hands of God (Box 1). Participants described waiting for severe symptoms before making decisions to seek care and limited knowledge of causes and course of illness was evident. A fear of Ebola was also reported as a deterrent from visiting health facilities during the outbreak. In general, participants recognised that Ebola was over and reported that it was no longer a concern.

### Delays in reaching healthcare facilities

IDI participants in rural areas reported that accessing care was a challenge. Reasons cited were lack of transport, cost of transport, and bad or unpassable roads, particularly during the rainy season. In rural health facilities, referrals were described as slow and contributed to delays in reaching appropriate care. Slow referrals were reported caused by transportation issues (a lack of available transport, combined with poor ambulance coverage and slow response time), as well as a lack of expertise among health staff in rural areas who did not make referrals until the situation became critical, often when it was too late (Box 2). HCWs also reported referral delays due to carers being unable to take decisions on referrals alone due to the financial implications, and/or feeling concerned about leaving their families (Box 2).

**Box 2:**
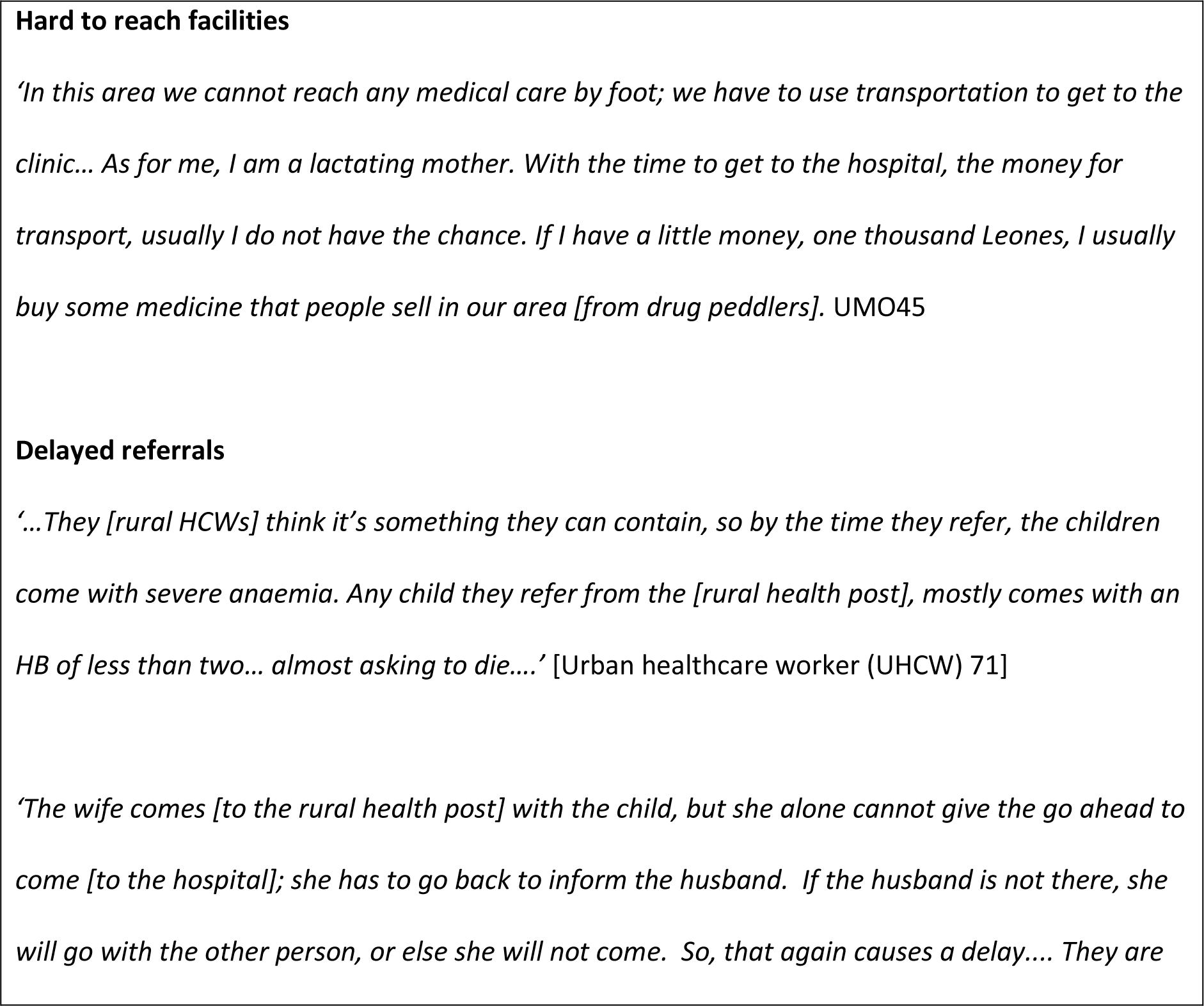
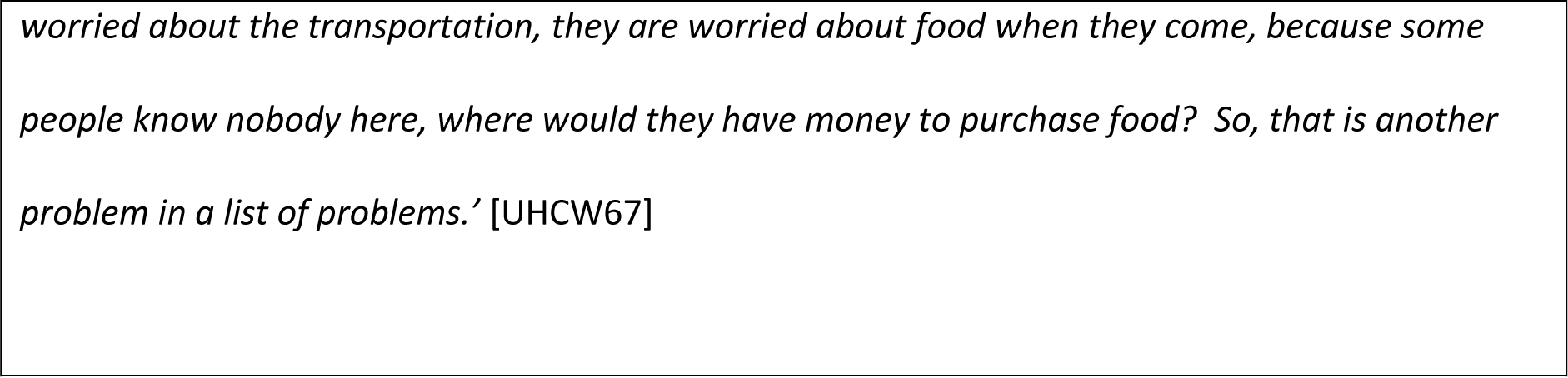
Themes and quotes relating to delays in reaching health facilities.

### Places where healthcare was sought and care preferences

Of those who stated that they sought advice/ treatment for a febrile child under 5 years, 90% reported that they sought care at a health facility at least once during the illness (Table 3). Of those who reported that they attended a health facility, 70% (297/425) of urban children vs 0.5% (2/365) of rural children attended a hospital. Carers in both areas also sought advice/ treatment from alternatives including pharmacies, shop/markets, state (official) community health workers (CHWs), and ‘drug peddlers’ (persons selling medications unofficially/ privately) (Table 3). Use of traditional healers was only reported by 2 (0.4%) urban carers but by 37 (9.2%) rurally (Table 3). The formal health sector (healthcare facility or state CHW) was reported as the first pattern of resort for 88% of urban and 86% of rural carers who did seek treatment and advice for the febrile child; and whilst no urban carers reported going first to a traditional healer, 8.5% rural carers did so (Table 3).

In contrast to the findings of the survey, IDI participants explained that due to the numerous barriers to accessing health facility based care they often sought care from an alternative provider first: generally a local pharmacy or off-duty nurse in the urban area, and a traditional practitioner or drug peddler in the rural area (Box 3). As a result, seeking care from the formal sector and health facilities was typically delayed until initial treatment was deemed to be ineffective and/or when symptoms were perceived to be severe (usually described as convulsions or acute anaemia) (Box 3).

**Box 3:**
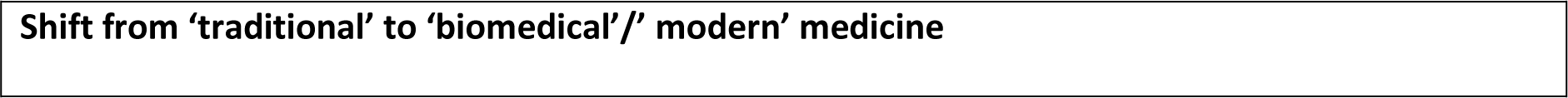
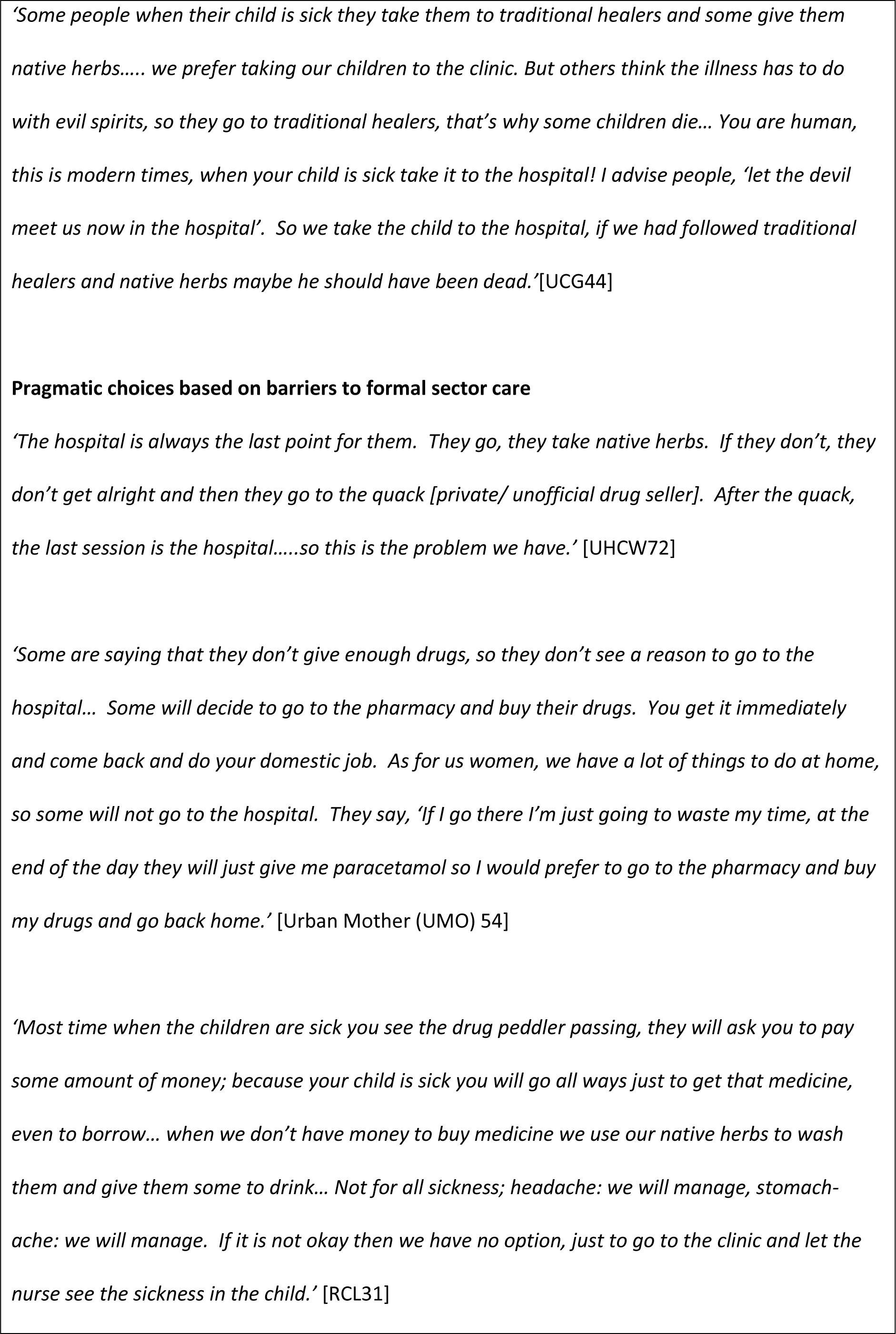
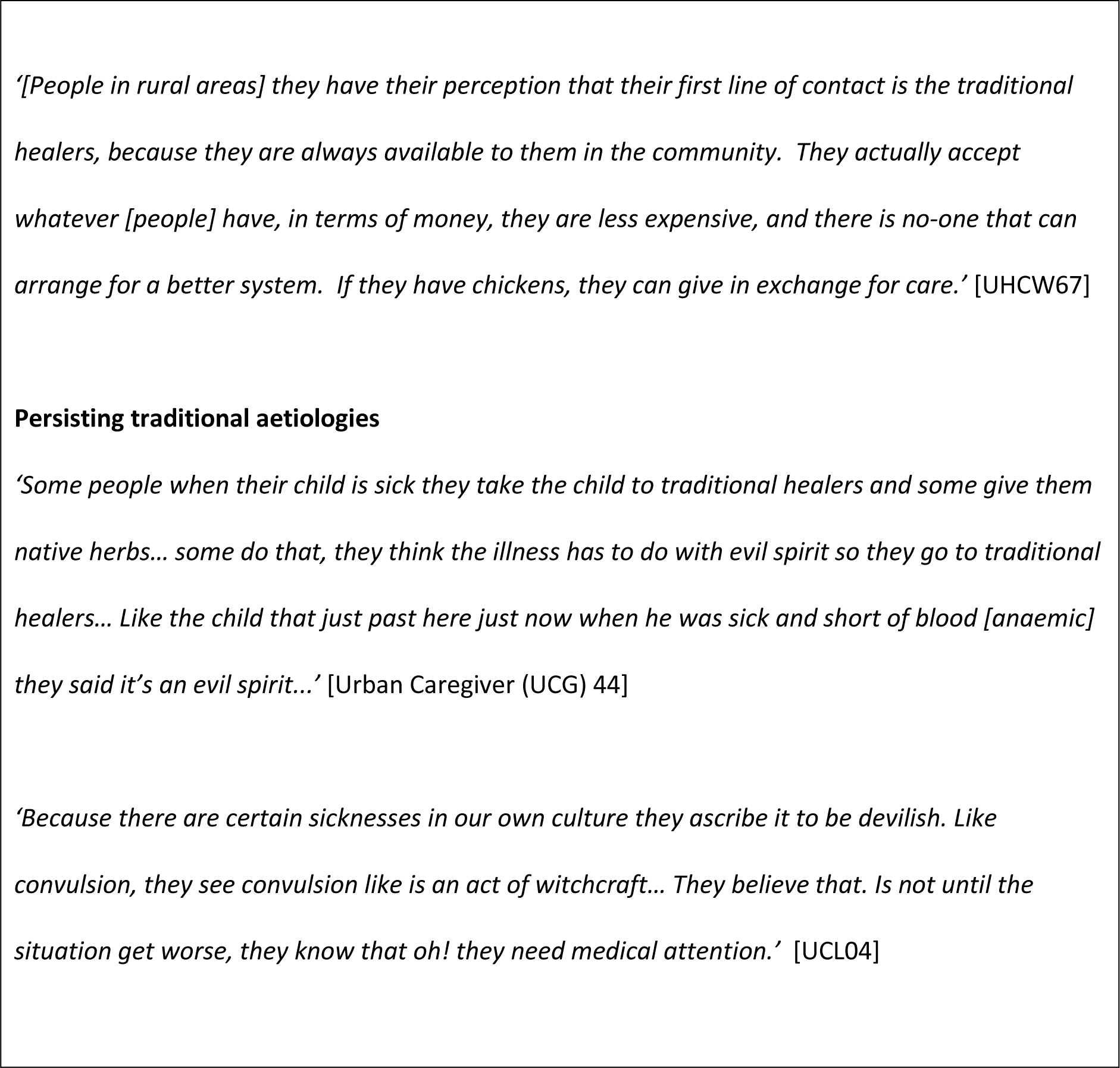
Themes and quotes relating to care preferences.

In line with survey findings, IDI participants described a general preference for formal sector care. Moreover, they explained a shift in practice over time as people increasingly sought care in health facilities instead of from traditional providers, and perceived ‘modern medicine’ to offer the best care for their children. Whilst seeking care from a traditional practitioner was generally pragmatic, some participants perceived that there were certain conditions for which traditional treatment was required (Box 3). Some participants sought herbal or spiritual remedies when biomedical solutions failed (Box 3).

### Healthcare experience and delays in receiving quality care

Of those who reached heath facilities for febrile illness, 95% of urban and 90% of rural children under 5 years received a blood spot test (usually indicative of having rapid malaria test) (Table 3). Whilst very few carers paid for this in the urban area, one third of rural carers were required to pay the HCW for the test (Table 3). Similarly, 94% of children attending health facilities for febrile illness received medications, though 5% urban vs 41% rural carers were required to pay for them (Table 3).

In addition to paying for medication and treatment, IDI participants explained routine charging for other aspects of care such as under-5 vaccination cards. In the rural area they also described they felt an obligation to ‘show a sign of appreciation’ to HCWs (giving money or food was perceived to be necessary to ensure good care in the future); in the urban area paying to be seen quickly was described as commonplace.

Several HCWs confirmed the practice of charging for care and explained this was necessary to compensate for their limited or non-existent salaries, inconsistent drug supplies, and a lack of other basic equipment (Box 4). Confusion about medications that should be ‘free’ and those that should be ‘cost recovery’, and why payment was required for certain drugs, generated mistrust between HCWs and communities (Box 4).

**Box 4:**
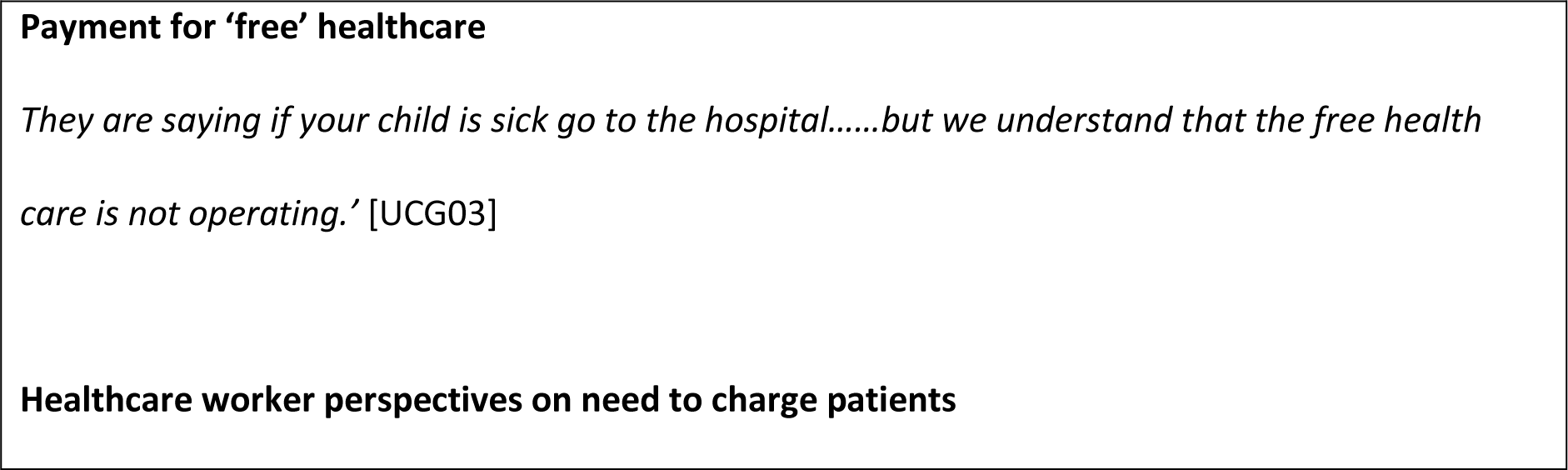
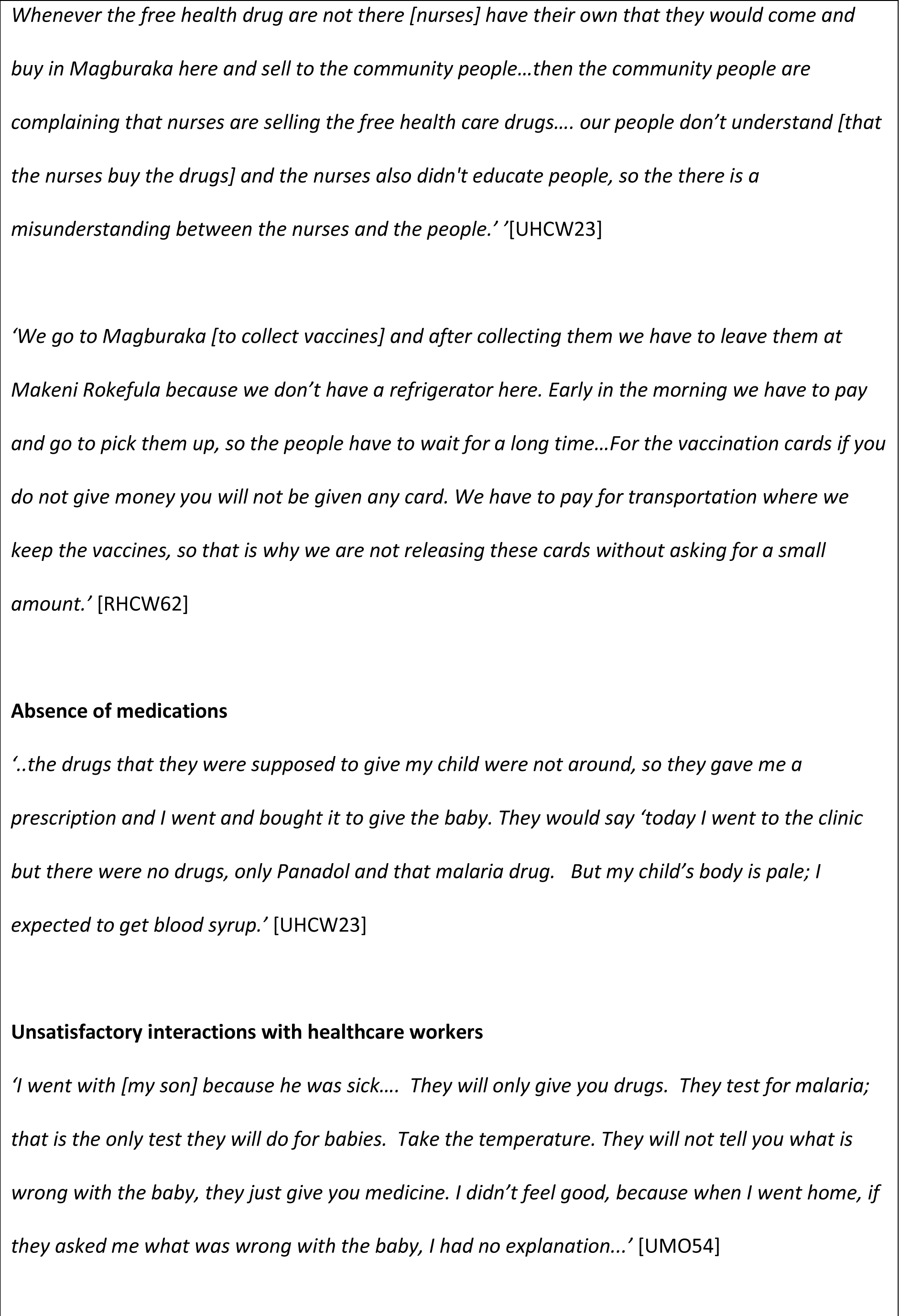
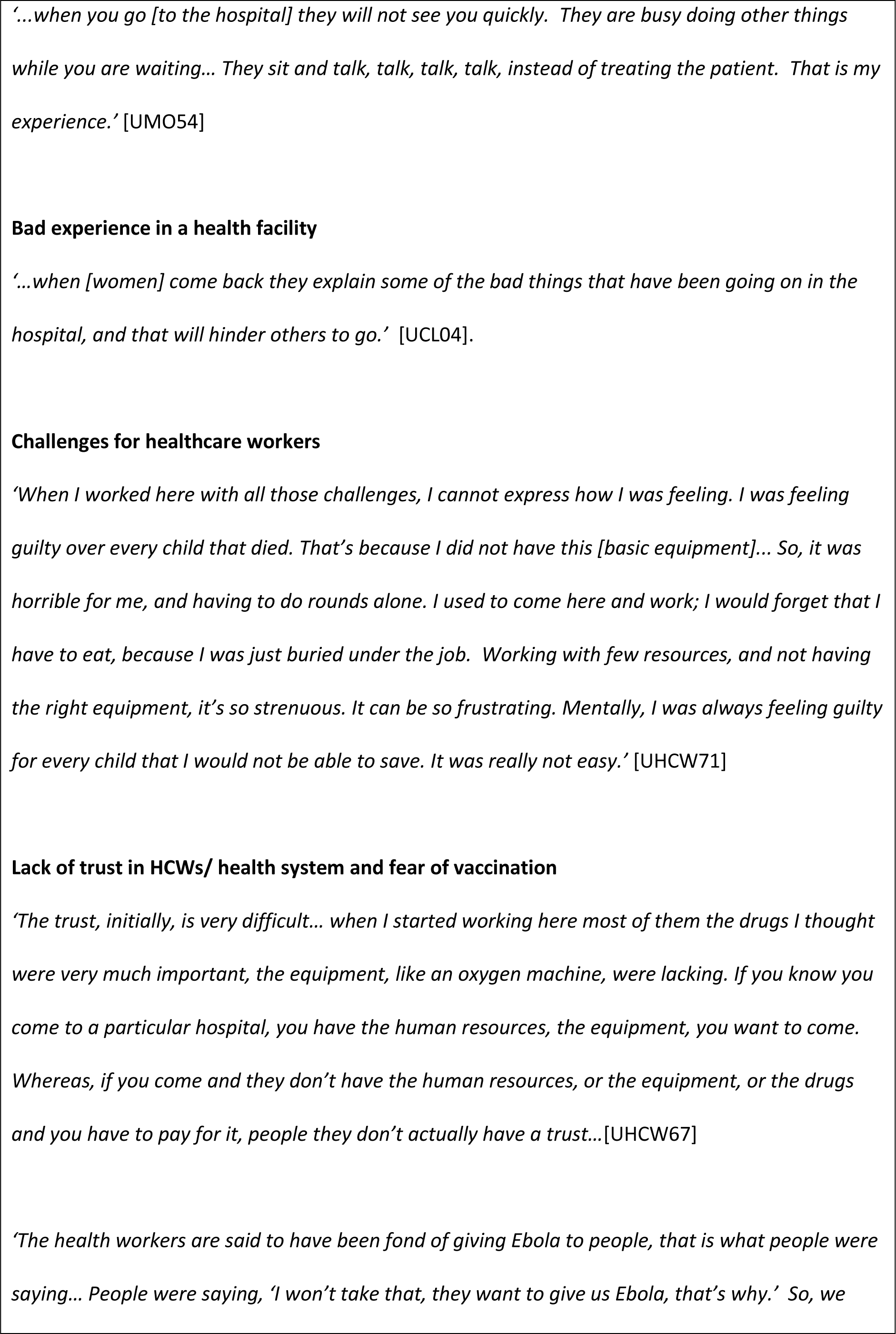
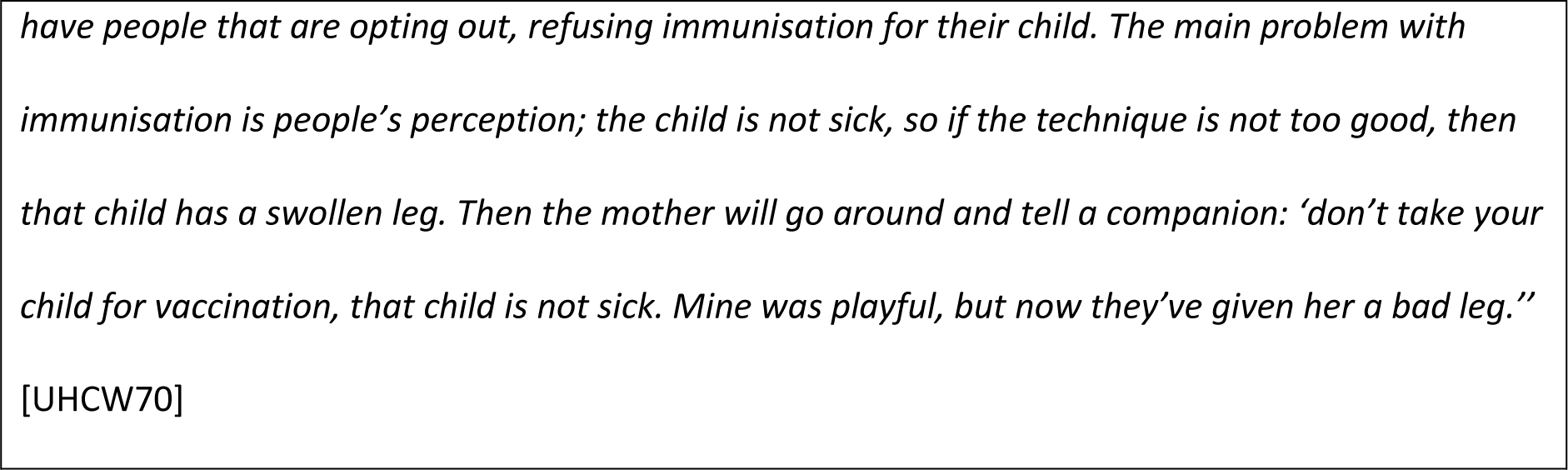
Themes and quotes relating to payment for healthcare, healthcare experience and quality, and vaccination.

Among IDI participants, rural mothers and carers in particular reported that HCWs were absent from the facility when they visited. In both urban and rural, poor quality of care provision by healthcare staff was reported as an important barrier to quality care. Carers felt communication by HCWs was poor; from inadequate explanations of diagnosis and care to experiences of disrespectful, aggressive and insulting treatment (Box 4). In some instances, rural participants explained this had led to a rift between the community and clinic nurse, to the extent that some would refuse to attend the local clinic (Box 4).

Some HCWs acknowledged they could treat patients harshly but explained this was due to their frustration working with ‘stubborn’ communities and the challenges of working with limited resources (poor infrastructure, lacking equipment, and interrupted drug supplies). They also described working in such conditions as negatively impacting their wellbeing and mental health (Box 4).

All IDI participant groups emphasised that they would not return to a health facility in which they had a bad experience, and instead would look for alternatives. Furthermore, such experiences were described to have a ‘multiplier effect’, as both bad and good experiences influence others’ decisions (Box 4).

### Vaccination and bed net coverage and barriers to vaccination

Vaccination cards were available for 54% of children whose carers were surveyed (Table 3). Amongst those eligible for the measles vaccine, at least one dose had been received by 86% in urban areas compared with 83% in rural areas (evidenced by vaccination card or disclosed by carer) (Table 3).

Vaccine coverage for at least one dose polio vaccine was 98% and 90% in urban and rural areas respectively (Table 3). Rurally, carers caring for more children were less likely to have vaccinated children (Table 4). Of those vaccinated for measles, 64% (274/425) of children in the urban areas and 45% (185/411) of children in the rural areas had been vaccinated during a national mass measles vaccination campaign conducted in May and June, 2016. Of those vaccinated for polio, 94% (488/520) of urban and 93% (469/504) of rural children had been vaccinated during the period of a national mass polio vaccination campaign (September and October 2016).

Overall, 25% (95%CI: 17-35) of urban carers vs 75% (95%CI: 60-84) of rural carers reported one or more barrier to getting their children vaccinated (Figure 2).

**Figure 2:**
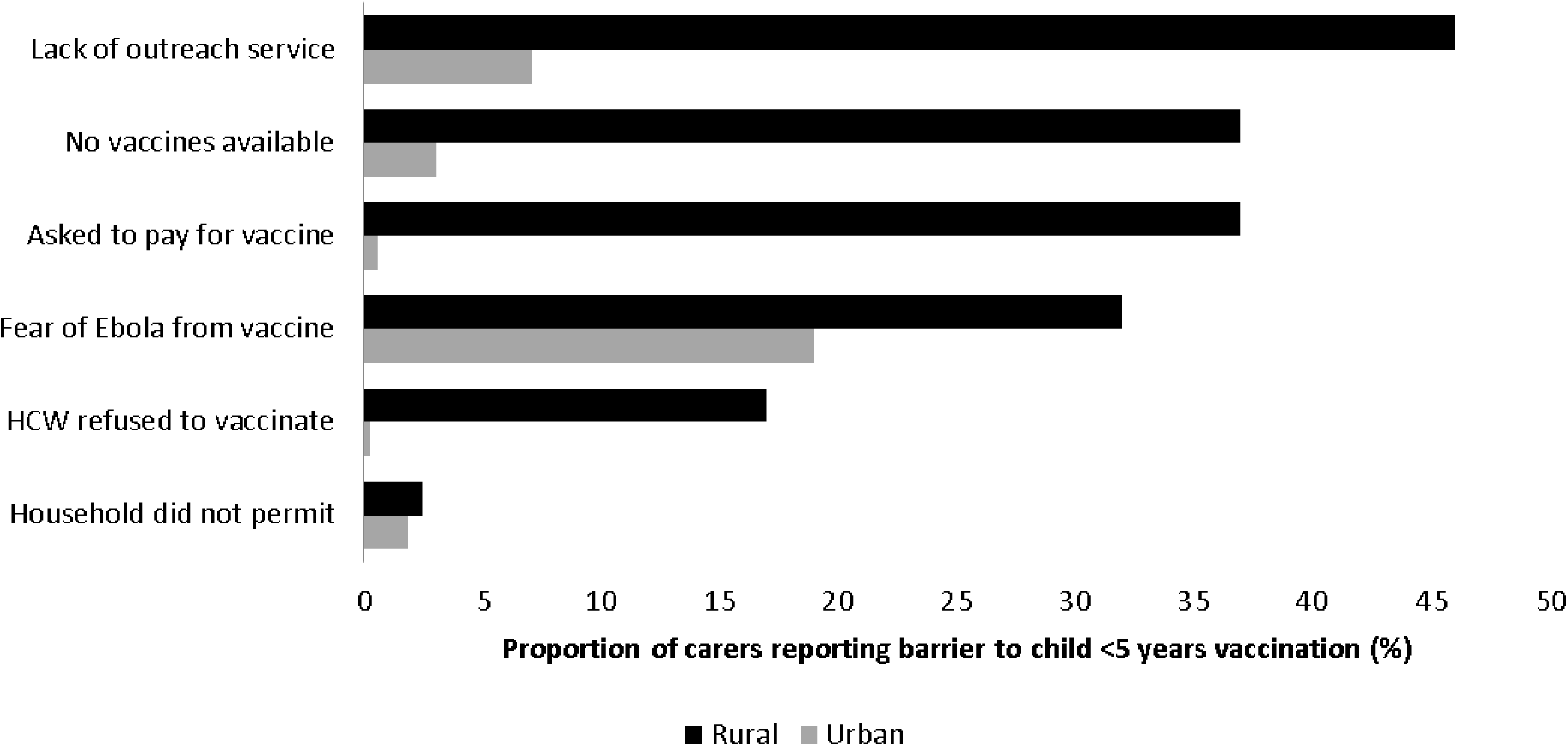
Barriers to receiving vaccination for children <5 years according to their carer, by place

Generally, IDI participants expressed a high level of vaccine acceptance and adherence. However, participants also described cost, distance to health facilities, lack of vaccines and issues relating to cold chain (e.g. lack of refrigerators at facilities and or cold boxes to transport vaccines) as major barriers to vaccination. HCWs explained that the same barriers also compelled them to charge for vaccines and vaccination cards, as they sought recompense for costs incurred collecting and storing vaccines (Box 4). Fear of Ebola had also impacted vaccination acceptance, according to HWCs and community leaders, but this was thought to be improving. There were concerns about vaccine side effects. HCWs and community leaders reported some hesitancy from communities and that some mothers had complained about the aftereffects of immunisation, notably rashes and bumps or abscesses (Box 4). This was particularly evident in areas where there were concerns about vaccine supply and the integrity of the cold chain.

When exploring bednets coverage, 31% of urban and 12% of rural households reported they possessed intact LLITNs. Around one quarter of urban and less than one tenth of rural children under 5 years and pregnant women reported sleeping under a LLITN the night before the survey (during rainy season) (Table 3). IDI participants described bednet distributions as insufficient and inequitable. Some community leaders also felt that of the importance of bednets was not recognised and improved awareness was required to decrease malaria rates.

## Discussion

This study has documented poor health indicators for children under 5years and high incidence of children febrile illness. This was particularly severe in rural areas, where there was a high prevalence of morbidity and the under 5 years mortality rate was close to the humanitarian emergency threshold (2.0 per 10,000/ day).[23] Although previous studies have demonstrated poor health indicators pre- and peri-Ebola, to our knowledge this is the first to demonstrate the extent of it, capturing burden, experiences and perspectives in the post Ebola period.

In terms of health behaviour, most carers in both areas sought care for febrile children under 5 years. However, substantial barriers delayed and prevented a large majority from obtaining healthcare when required. Decisions on seeking care were often made hierarchically or collectively, often entailing delay. Women in particular were obliged to defer to men or elder family members. The decision to seek care for children was often delayed until a child was seriously unwell in view in anticipation of perceived barriers to healthcare, particularly distance and financial costs. In other words, critical delays in deciding to seek care (first aspect of delay model) was strongly influenced by the fear/anticipation of practical problems faced in reaching a health facility (second aspect of delay model) and receiving adequate and appropriate healthcare (delay phase 3). Participants linked delays and aborted attempts to reach healthcare facilities resulting from barriers with severe consequences including death.

There was an expressed preference for choosing biomedical healthcare for unwell children over traditional methods and alternatives, particularly in the urban area. Though IDI findings suggest that traditional and use of alternative providers was probably more common than reported by the survey, biomedical healthcare was generally considered safer and more effective than alternatives. This is a salient finding and runs contrary to prior reports and widely held perceptions that people prefer traditional therapies.[24] People generally chose alternatives to biomedical healthcare as informal care providers and traditional healers were often considered more convenient, accessible and affordable. However, beliefs in traditional means to treat certain conditions/presentations were also apparent.

Whilst most children under 5 years who reached healthcare facilities received investigation and medication, for almost a half of rural carers payment for “free healthcare” was necessary. Payment was less commonly reported in the survey in the urban areas though IDI findings suggest payment is commonplace in both areas including for vaccinations. Lack or absence of HCWs, and lack of medications were commonly reported. Poor relations and lack of trust between people and HCWs was evident and largely attributed to payment of HCWs and suspicion of personal gain from the sale of ‘free healthcare’ and exacerbated by suboptimal interactions with HCWs. However, our study has also provided HCW perspectives and offers insight into factors leading to apparent ‘corruption’ and suboptimal provision of care. HCWs often described low or absent pay, difficult working conditions, a lack of support. HCWs would seek compensation/payment from patients to recover routine costs that they incurred personally. Issues of poor referral mechanisms and a lack of ambulances were also evident. Our findings indicate inequity in healthcare access and inequality in health between urban and rural areas. There was also a probable socio-economic divide within areas with more educated carers and those caring for fewer children more likely to have sought care for febrile children and the child to have received vaccines.

Measles and polio vaccination coverage was higher than previous surveys though was below the threshold for herd immunity for measles.[25] The higher than anticipated vaccination coverage probably reflects relatively successful mass vaccination campaigns. Indeed, evidence from this study suggests routine vaccination services were poor, subject to routine charging, and that there were persistent issues with vaccine hesitancy. Very poor LLITN coverage is also supported by the high proportion of children experiencing febrile illness compatible with malaria during the study recall period. Recommended treatment is, in theory, available free to all through the Sierra Leone malaria program, but delays and lack of access to such treatment was reported by the majority. This will impact long-term morbidity and mortality due to malaria. Lack of bednet use by the majority means at least one of the most effective preventive interventions was not systematically implemented.

This study draws on complementary strengths of using mixed methods. Preliminary survey analysis revealed several of the poll’s shortcomings and potentially disputed conclusions, which had a direct impact on the design and execution of the in-depth interview study. As a result, we were able to spot trends and discrepancies in the data, which let us come to solid conclusions in which we have a high degree of confidence.

This study is subject to limitations. For selection of rural clusters, mapping was not practical and limitations with rural population estimates precluded both probability sampling proportional to size and weighting in the analysis. Smaller village residents might have been overrepresented in the study, which could have inflated the discrepancies between the two areas. Although it’s likely that some respondents choose not to declare their eligibility, the high participation percentage is comparable to earlier polls.[25] The high participation rate is similar to previous surveys, though it is possible that some participants declined to declare eligibility.[25]

With individuals perhaps under-reporting socially unfavourable actions, some responder bias is expected; as such we have probably over-estimated facility-based healthcare seeking for febrile children in the survey. However, IDI participants also commonly reported use of alternatives to healthcare. Similarly, we are likely to have over-estimated vaccination coverage especially given we were reliant on a verbal report in many cases. Under-reporting of child deaths is also possible with mothers and carers fearing blame, judgement and experiencing feelings of guilt especially if healthcare was not sought. The period of recall was long for child mortality and the time of death may have been challenging to recall. For child illness the recall was relatively short (three months) but for a carer with multiple children the last episode of febrile illness may be difficult to recall.

Regarding the generalisability of the results, it is acknowledged that this study was conducted in a relatively small geographical area and there are variations in the socio-economic makeup and availability of health services between districts, with Tonkolili’s residents being among the poorest and underserved. Nevertheless, Yoni is thought to be comparable to many areas of rural Sierra Leone by MoHS and MSF. MSF supports the hospital in Magburaka by providing resources, staff, and training so that urban residents who live in Magbruaka or nearby may have easier access to free healthcare than populations of a similar size in other districts.

Findings have recently been corroborated by a qualitative study identifying an exacerbation of existing barriers on delivery of bed net and immunisation programme due to Covid-19.[26]

### Conclusions

Our study indicated that children under 5 years encounter significant barriers to care, especially in rural areas, resulting in high preventable morbidity and mortality near the emergency threshold rurally. People want to access healthcare, but available services are often costly (despite the national policy for free care), unreachable, and poor quality; on the other hand, lack of pay and holistic support precluded healthcare workers to deliver quality of care. Practical actions are required to address barriers to reach and deliver care and reduce preventable deaths.

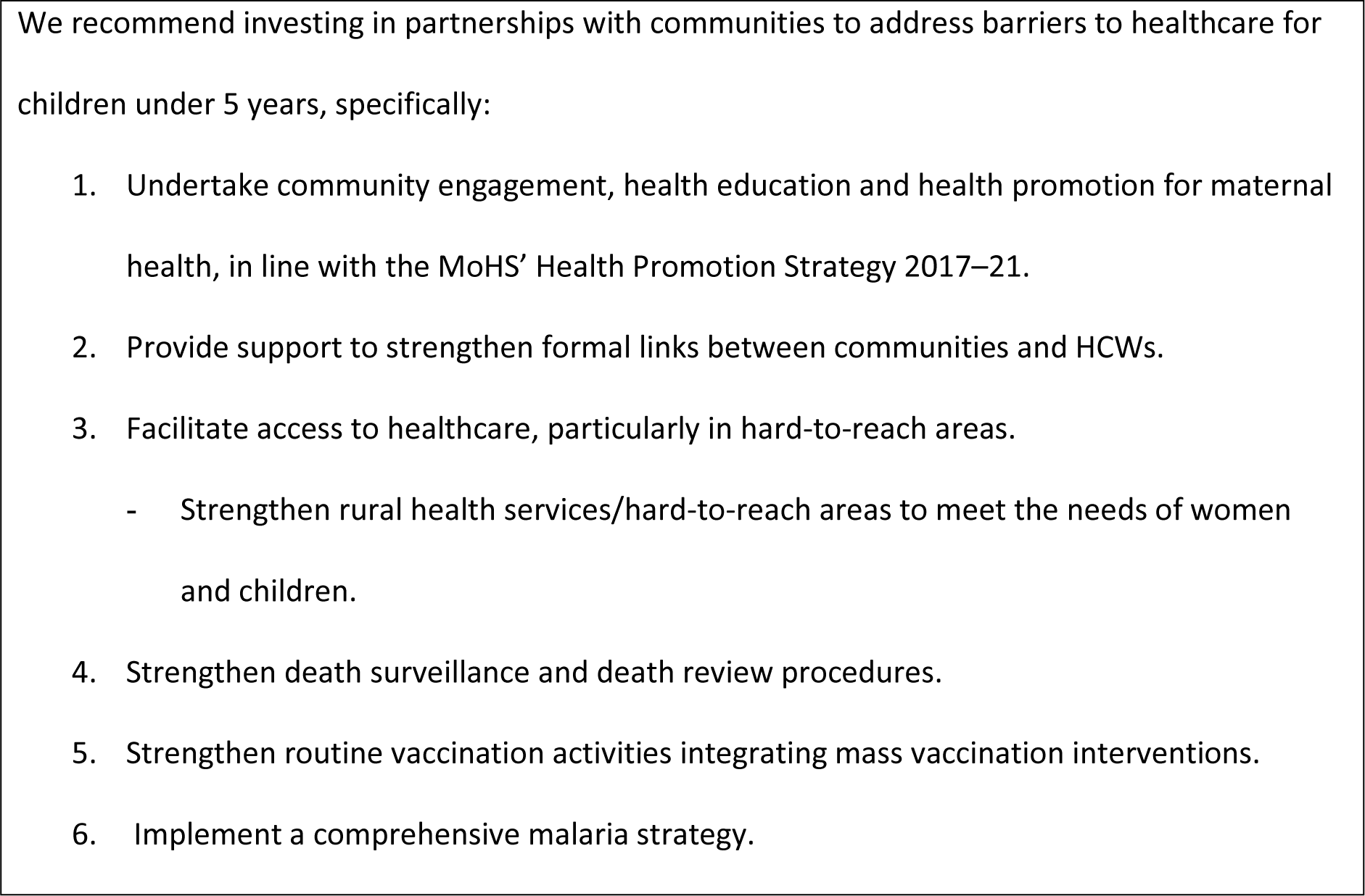

## Data Availability

All data produced in the present study are available upon reasonable request to the authors.

## Acknowledgements

This work would not have been possible without support from the Sierra Leone Ministry of Health and Sanitation. With thanks to Antonio Isidro Carrion Martin, Pete Masters, Rosamund Southgate, Barbara Nasto, Idriss Ait-Bouziad, Paul Stewart, Kiran Jobanputra and Jonathan Mazliah; the MSF team in Tonkolili; the UK Field Epidemiology Training Programme (FETP); and the European Programme for Intervention Epidemiology Training (EPIET). Finally, we would like to acknowledge the dedication of the teams in Tonkolili as well as the people of Tonkolili who contributed their valuable time.

## Author contributions

JE implemented the study and wrote the first draft of the manuscript; KD implemented and supported data analysis; NG implemented and lead the qualitative data analysis; KW implemented and supported qualitative data analysis; BB supported study conceptualisation; BS supported design of qualitative methods; AJ, AB, and MS supported study design and implementation; KL, DK, and SS supported study conceptualisation; HB supported manuscript development; CS supported implementation; GC supported study conceptualisation and coordination. All authors contributed to first and last versions of the manuscript.

## Ethics and consent statement

The study protocol can be found at https://remit.oca.msf.org/studies/141. The protocol was approved by the MSF Ethics Review Board and the ethics committee of the Ministry of Health and Sanitation of Sierra Leone. Approval to was granted from traditional authorities in all proposed sites approved the study. Local languages of Sierra Leone (Temne, Mende or Krio) are not written languages, therefore – out of necessity – informed verbal witnessed consent was obtained from the head of the household and the parent/guardian in the selected household. In the case of younger participants who were between 15 and 17 years of age, permission of the parents or legal guardians was sought as both the legal age of consent and the minimum age of marriage is 18 years old.

## Paper context

### Main findings

In Tonkolili District, Sierra Leone, children under 5 years encountered significant barriers to care, especially in rural areas, resulting in high preventable morbidity and mortality. There was inequity in healthcare access and inequality in health between urban and rural areas.

### Added knowledge

This study documents poor health indicators for children under 5years and high incidence of children febrile illness, particularly severe in rural areas, where the under 5 years mortality rate was close to the humanitarian emergency threshold.

### Global health impact for policy and action

This study explores barriers to healthcare services and makes recommendations for policy and practice in Sierra Leone and similar contexts, specifically, investing in partnerships with communities to address barriers to healthcare for children under 5 years.

